# Pre-infection COVID-19 vaccination and long-COVID mental fatigue severity: Findings from the Johns Hopkins COVID Long Study

**DOI:** 10.1101/2025.11.07.25339684

**Authors:** Madeline Sagona, Zhanmo Ni, Eryka Wentz, Karine Yenokyan, Thea Kammerling, Andrea DeVito, Priya Duggal, Shruti H. Mehta, Bryan Lau

## Abstract

**Background:** Long-COVID is a post-acute sequela of SARS-CoV-2 infection characterized by persistent, multi-system symptoms. Neurologic symptoms, such as mental fatigue, are often reported. While vaccination prior to infection is known to lessen symptom burden, its impact on mental fatigue remains unclear.

**Objective:** Examine the association between vaccination and long-COVID-associated mental fatigue.

**Methods:** We analyzed data from the Johns Hopkins COVID Long Study, a cohort study of 22,811 participants with and without infection. Among 2,634 participants with complete longitudinal follow-up, we examined the association between pre-infection vaccination and mental fatigue as measured by the Wood Mental Fatigue Inventory (WMFI; range 0-36, with lower scores indicating less fatigue). We considered three groups: long-COVID, recovered, and never infected, with the latter two groups as negative controls. We estimated the score differences between vaccinated and unvaccinated participants using covariate-adjusted quantile regression and mixed-effects linear models.

**Results:** Among participants with long-COVID, receiving a booster dose was associated with lower WMFI scores across the distribution (1.5, 2.6, and 3.8 points lower at the 25^th^, 50^th^, and 75^th^ percentiles; *p* ≤ 0.02) compared to those unvaccinated. Fully vaccinated participants also had lower scores, though these differences were not statistically significant. Findings were consistent in mixed-effect linear models where boosted (4.0 points lower) and fully vaccinated (1.9 points lower) participants with long-COVID had lower WMFI scores (*p* < 0.05). No associations were observed among recovered or never-infected participants.

**Conclusion:** Pre-infection vaccination was associated with less long-COVID-associated mental fatigue, with the greatest benefit among boosted participants.

## Introduction

Severe acute respiratory syndrome coronavirus 2 (SARS-CoV-2) can lead to post-acute sequelae characterized by chronic symptoms and long-term disability in a subset of individuals [1]. The World Health Organization (WHO) defines SARS-CoV-2 post-acute infection syndrome (often termed “long-COVID”) as new or continuing symptoms three months after initial infection that last at least two months and cannot be otherwise explained [2]. Recent studies estimate that the prevalence of long-COVID is between 5.3% and 15% [3–6], although the true burden continues to evolve alongside the SARS-CoV-2 virus [7]. Long-COVID encompasses an array of debilitating symptoms including unremitting physical and mental fatigue, cognitive impairment, post-exertional malaise, autonomic dysfunction, and gastrointestinal symptoms [8,9].

The toll of mental fatigue is particularly pernicious. Among those with long-COVID, mental fatigue is one of the most common and debilitating symptoms [10]. Marked by a profound and pervasive sense of mental exhaustion, mental fatigue can impair cognitive [11] and emotional [12] functioning, limiting an individual’s ability to perform everyday tasks. Mental fatigue has also been associated with longer reaction times and higher rates of car accidents [13–15] and is often characterized by difficulty concentrating, memory problems, slowed thinking, and struggles with decision-making and word retrieval [16].

Observational studies and meta-analyses indicate that SARS-CoV-2 vaccination prior to infection lowers the risk of developing long-COVID relative to no vaccination. Two large studies utilizing data from the United States (US) Department of Veterans Affairs found that individuals vaccinated pre-infection had a lower cumulative incidence of long-COVID at one-year post-infection [7], and those who received both initial mRNA doses pre-infection were less like to receive long-COVID care [17]. Similarly, in a retrospective analysis of over 240,000 infected patients, those who received at least one vaccine dose were less likely to report two or more long-COVID symptoms [18]. Additionally, a survey-based study of over 3,000 participants from the United Kingdom (UK) found that double-vaccinated participants had a lower incidence of long-COVID compared to unvaccinated matched controls [19]. Meta-analyses further corroborate these findings, showing lower odds of developing long-COVID after one or two vaccine doses relative to no vaccination [20,21].

In addition to lowering incidence, emerging evidence indicates that vaccination may also attenuate the severity of persistent long-COVID symptoms. In a multisite study of 3,663 participants, Gottlieb et al. [22] found that those who were vaccinated prior to a long-COVID diagnosis reported fewer or less severe physical and mental health symptoms than unvaccinated individuals. Maier et al. [23] suggested that pre-infection vaccination reduces the total number of long-term symptoms, and that these symptoms were, on average, less severe compared to unvaccinated individuals.

However, it remains unclear whether vaccination attenuates the burden of mental fatigue in individuals with long-COVID. The extent to which pre-infection vaccination status relates to mental fatigue severity remains poorly characterized. Clarifying this relationship could inform clinical care beyond infection protection.

In this study, we examined the association between pre-SARS-CoV-2 infection vaccination status and long-COVID-associated mental fatigue in a nationwide, virtual cohort study, the Johns Hopkins COVID Long Study (JHCLS). To assess whether this relationship is specific to long-COVID, we also evaluated the association between vaccination status and mental fatigue in two groups serving as negative controls: participants who recovered from COVID-19 without persistent symptoms (“recovered-COVID”) and participants who were never infected (“never-infected”).

## Methods

The JHCLS is comprised of individuals with and without a self-reported history of SARS-CoV-2 infection; participants are asked to complete a one-time, short online survey at baseline with the option to consent to longitudinal follow-up every 3 or 6 months. Recruitment, study design, enrollment, and baseline demographics have been previously described [24]. The Johns Hopkins Bloomberg School of Public Health Institutional Review Board (IRB) reviewed and approved this study on December 15, 2020.

### Case definitions

Long-COVID status was defined using the WHO definition [2]: participants who had at least 12 weeks between their initial SARS-CoV-2 infection and baseline survey completion and reported at least one new or continuing symptom were defined as having long-COVID. Participants who had at least 12 weeks between initial infection and baseline but did not report any new or continuing symptoms were defined as having recovered-COVID. A subset of participants reported at least one new or continuing symptom at baseline but had fewer than 12 weeks between infection and baseline survey completion. For these participants, we assessed their first follow-up visit to determine whether they experienced any new or continuing COVID-19 symptoms since baseline. If this was the case, they were also classified as long-COVID. Participants without a history of infection were classified as never-infected.

To determine perceived mental fatigue severity, we utilized the Wood Mental Fatigue Inventory (WMFI) [16], a short questionnaire of nine domains (scored 0-4) for a range of 0-36, with higher scores indicating greater mental fatigue burden. To contextualize the range of WMFI scores, we used the 90^th^ percentile of scores in the never-infected group to determine a cut point of mental fatigue disability, or severe interference in daily life, and this was set at 20. A score of 30-34 represented severe mental fatigue disability, and a score of 35-36 represented critically severe mental fatigue disability [25].

The WMFI was originally validated in a study [16] comparing non-clinical subjects with professions and/or social groups likely to experience fatigue (e.g., nursing) with four clinical groups: patients with chronic fatigue syndrome (CFS), patients with recovered CFS, patients experiencing depression, and patients with muscular dystrophy. Respondents were asked how much they were bothered by nine domains of mental fatigue over the past month (0 - “not at all”, 1 - “a little”, 2 - “somewhat”, 3 - “quite a lot”, or 4 - “very much”). For the purpose of the JHCLS, participants were asked to rate how much they were bothered by these nine domains over the past two weeks. The WMFI was found to have high internal consistency (Cronbach’s α = 0.93) and good test–retest reliability among the non-clinical group who completed the questionnaire twice (Pearson’s *r* = 0.887) [16].

### Cohort

For this analysis, we identified three groups of participants: long-COVID, recovered-COVID, and never-infected. For participants with a history of infection, we utilized baseline data on SARS-CoV-2 infection, vaccination history, sociodemographic factors, and clinical characteristics, along with longitudinal mental fatigue assessments. As of the data freeze on October 23, 2023, 21,675 participants with a history of infection provided informed consent to participate in baseline (Fig. 1). Of those, 3,998 also provided informed consent to participate in longitudinal follow-up every three months. We included participants who completed at least one WMFI assessment during follow-up, had a defined long-COVID status, and were likely infected during the months when the Alpha (February - June 2021), Delta (July - November 2021), or Omicron variant (December 2021 onward) was dominant in the US [26]. After excluding international participants due to small sample size (*n* = 134), participants with invalid or missing vaccination dates (*n* = 74), and participants infected with the “ancestral” strain (*n* = 1,440), the final population consisted of 1,734 participants. We excluded participants infected with the ancestral strain to mitigate bias due to strong confounding between variant type and vaccination availability.

**Fig. 1.**
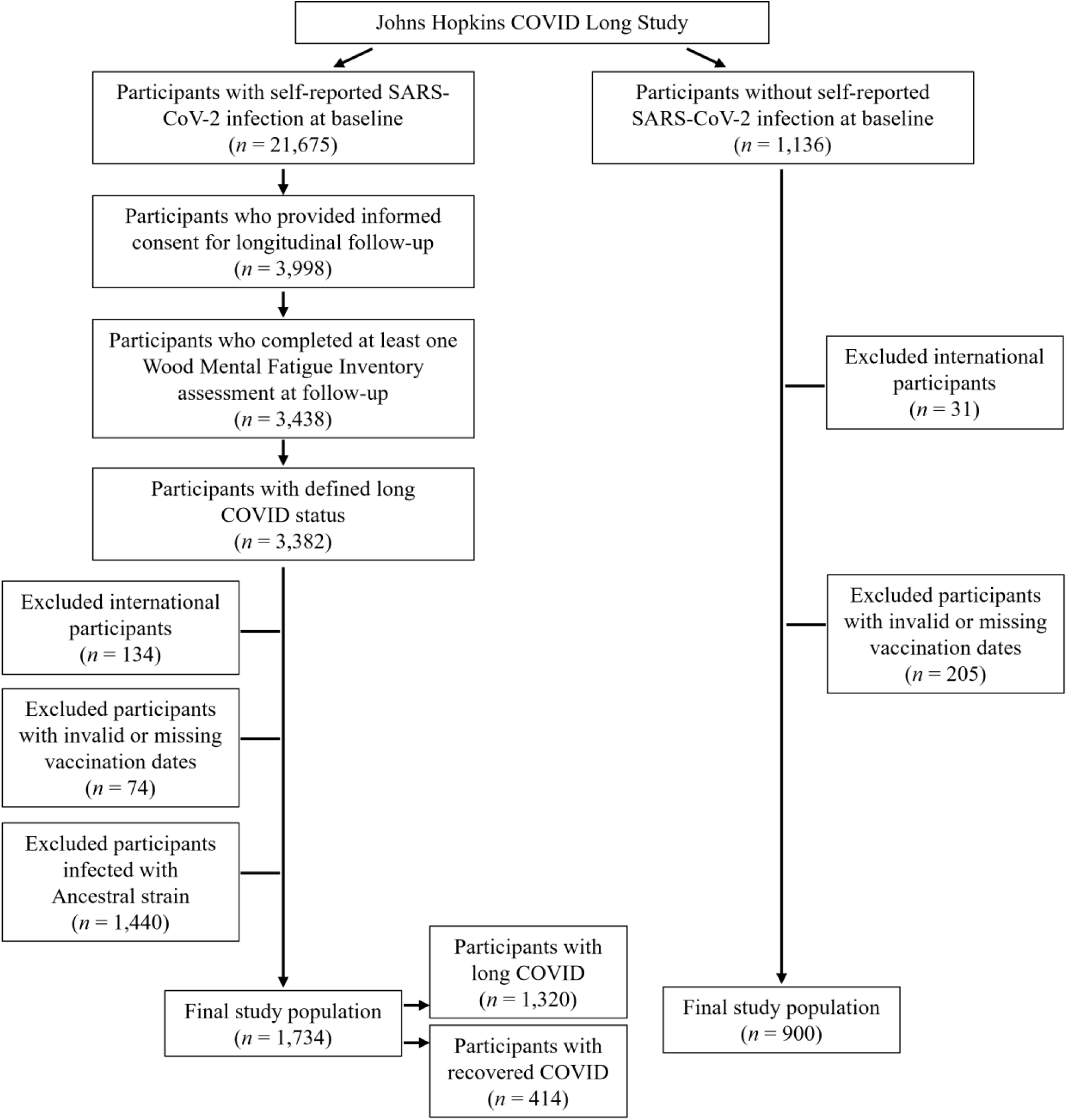
Flow chart of participant selection and distribution of long COVID status in final population.

For participants without a history of SARS-CoV-2 infection, we utilized baseline data on vaccination history, sociodemographic factors, clinical characteristics, and mental fatigue. As of the data freeze on November 10, 2023, 1,136 participants provided informed consent to participate in baseline. After excluding international participants due to small sample size (*n* = 31) and participants with invalid or missing vaccination dates (*n* = 205), the final population of participants without a history of infection was 900.

### Sociodemographic and Clinical Factors

Vaccination status was modeled as a categorical variable based on the number of COVID-19 vaccines reported prior to initial infection for those with a history of infection or at the time of the baseline survey for those without a history of infection. Participants were classified into one of four vaccination status groups: unvaccinated, partially vaccinated, fully vaccinated, or boosted. Participants reporting vaccination were classified as partially vaccinated if they reported one dose in a two-dose vaccine series, fully vaccinated if they reported a completed primary series (two doses in a two-dose vaccine series or one dose in a single-dose vaccine series), or boosted if they received at least one additional dose beyond the primary series. In descriptive analysis, vaccines were grouped according to vaccine platform.

For participants with a history of infection, we included time since infection as a continuous variable centered at 84 days post-infection such that the coefficient for time represented how WMFI score (continuous) changed after the 12-week mark (to align with the WHO’s definition of long-COVID). Interaction terms between vaccination status and time since infection were examined to test for modification effects. Analyses were adjusted for demographic covariates which included age (continuous), gender (men, women, different identity), and race (white, Black, East Asian, South Asian, Native American/American Indian/Alaska Native, Native Hawaiian/Pacific Islander, multiracial, other).

Socioeconomic and geographic variables included region of the US (Northeast, South, Midwest, West) and occupation (collapsed into 11 categories; S Table 2). Clinical variables were defined as conditions existing prior to initial infection or prior to the COVID-19 pandemic for those without a history of infection. These comorbidities included: diabetes, cardiovascular disease, chronic kidney disease, chronic lung disease, depression, anxiety/other mental health condition, history of stroke, overweight/obese, and asthma/reactive airway disease.

## Statistical Analysis

We evaluated the relationship between WMFI score and COVID-19 vaccination status, controlling for sociodemographic and clinical factors. Before fitting regression models with covariates, we examined the unadjusted distribution of WMFI scores across infection-status groups (never-infected, recovered-COVID, and long-COVID) (Fig. 2). To account for repeated measures without overweighting participants with more visits, we computed each participant’s mean WMFI across all available follow-up visits and used these participant-level means for group comparisons. We summarized group-level distributions with the mean, median, and interquartile range, and visualized them with boxplots.

**Fig. 2.**
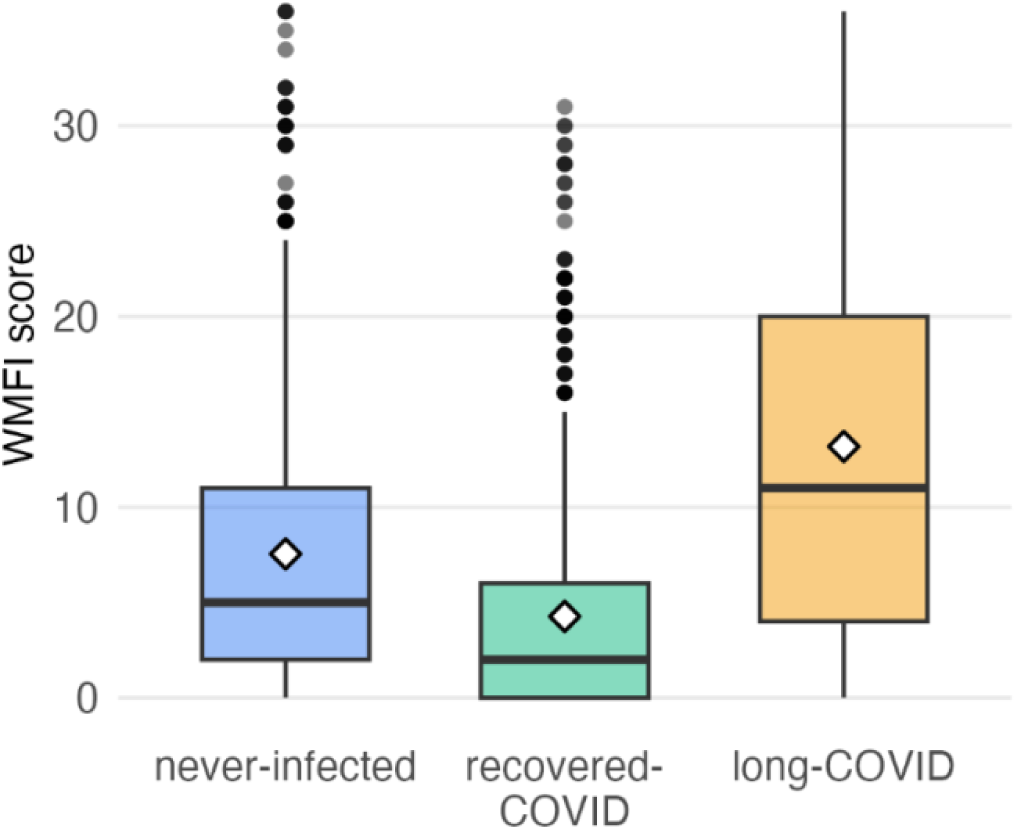
Wood Mental Fatigue Inventory score distribution by infection and long-COVID status.^a^. Abbreviations: WMFI: Wood Mental Fatigue Inventory, IQR: interquartile range. ^a^Diamonds indicate means.

We utilized quantile regression to account for the skewed distribution of WMFI scores. Quantile regression is a non-parametric approach that yields consistent estimators when there is dependence between repeated measurements [27,28]. To account for repeated WMFI measures among participants, 95% confidence intervals (CIs) were obtained through a cluster bootstrap [29–32]. We estimated the probability density function of the WMFI score by vaccination status using an Epanechnikov kernel function with bandwidth as the standard deviation of the smoothing kernel to a dense set of quantiles (0.05 to 0.95 in increments of 0.01). Never-infected and recovered-COVID participants were used as negative controls to confirm the absence of a significant association (see Fig S1 for directed acyclic graph) [33].

For completeness, we accompanied the quantile inferences with an analysis modeling the mean difference between those who were and were not vaccinated by mixed effect linear regression models with participant-level random intercepts. Models were fitted using restricted maximum likelihood estimation, which permits inference about the subject-specific mean of WMFI scores after adjusting for potential confounders.

Finally, to assess the robustness of our results across different SARS-CoV-2 variants, we conducted sensitivity analyses by performing the quantile regression and mixed effect models restricting to participants likely infected with the Omicron variant (December 2021 onward) to examine whether the associations held among participants infected during the most recent years of our data.

## Results

### Descriptive characteristics

A total of 2,634 participants were analyzed, including 1,734 participants with a history of SARS-CoV-2 infection (76% long-COVID, 24% recovered-COVID) and 900 without a history of infection (Table 1). The median age of participants was 46 years (interquartile range (IQR): 34–58). Overall, the cohort was predominantly female (83%) and white (87%), with nearly half residing in the southern US (45%), and half reporting any post-graduate studies (51%). Participants with long-COVID contributed a median of three follow-up visits (IQR: 3–5). Among all participants, 15% were unvaccinated, 2.4% were partially vaccinated, 24% were fully vaccinated, and 59% were boosted. Among vaccinated participants, mRNA vaccines were the most commonly reported vaccine platform (>90% across all groups).

**Table 1.** Sociodemographic and clinical characteristics of participants in the Johns Hopkins COVID Long Study (*n* = 2,634).

|  | Never-Infected<br>(*n* = 900) |  |  |  | Recovered-COVID<br>(*n* = 414) |  |  |  | Long-COVID<br>(*n* = 1,320) |  |  |  |
| --- | --- | --- | --- | --- | --- | --- | --- | --- | --- | --- | --- | --- |
|  | Unvaccinated<br>(*n* = 30) | Partially<br>Vaccinated<br>(*n* = 8) | Fully<br>Vaccinated<br>(*n* = 111) | Boosted<br>(*n* = 751) | Unvaccinated<br>(*n* = 31) | Partially<br>Vaccinated<br>(*n* = 7) | Fully<br>Vaccinated<br>(*n* = 117) | Boosted<br>(*n* = 259) | Unvaccinated<br>(*n* = 321) | Partially<br>Vaccinated<br>(*n* = 48) | Fully<br>Vaccinated<br>(*n* = 408) | Boosted<br>(*n* = 543) |
| **Age** (mean ± SD) | 44 (16) | 35 (9) | 42 (16) | 44 (16) | 50 (15) | 48 (16) | 42 (14) | 46 (16) | 48 (13) | 46 (13) | 46 (14) | 48 (15) |
| **Gender** |  |  |  |  |  |  |  |  |  |  |  |  |
| Men | 6 (20%) | 0 (0.0%) | 16 (14%) | 95 (13%) | 13 (42%) | 1 (14%) | 23 (20%) | 64 (25%) | 46 (14%) | 4 (8.3%) | 62 (15%) | 77 (14%) |
| Women | 23 (77%) | 7 (88%) | 92 (83%) | 636 (85%) | 18 (58%) | 6 (86%) | 94 (80%) | 193 (75%) | 274 (85%) | 43 (90%) | 342 (84%) | 452 (83%) |
| Different identity | 1 (3.3%) | 1 (13%) | 3 (2.7%) | 20 (2.7%) | 0 (0.0%) | 0 (0.0%) | 0 (0.0%) | 2 (0.8%) | 1 (0.3%) | 1 (2.1%) | 4 (1.0%) | 14 (2.6%) |
| **Race** |  |  |  |  |  |  |  |  |  |  |  |  |
| White | 27 (90%) | 6 (86%) | 88 (82%) | 609 (82%) | 25 (81%) | 6 (86%) | 102 (88%) | 225 (87%) | 294 (92%) | 45 (94%) | 352 (86%) | 501 (92%) |
| Black | 1 (3.3%) | 0 (0.0%) | 5 (4.6%) | 12 (1.6%) | 2 (6.5%) | 0 (0.0%) | 2 (1.7%) | 7 (2.7%) | 9 (2.8%) | 3 (6.2%) | 20 (4.9%) | 10 (1.8%) |
| East Asian | 0 (0.0%) | 0 (0.0%) | 5 (4.6%) | 46 (6.2%) | 2 (6.5%) | 1 (14%) | 3 (2.6%) | 6 (2.3%) | 2 (0.6%) | 0 (0.0%) | 1 (0.25) | 5 (0.9%) |
| South Asian | 1 (3.3%) | 0 (0.0%) | 4 (3.7%) | 28 (3.8%) | 0 (0.0%) | 0 (0.0%) | 0 (0.0%) | 6 (2.3%) | 2 (0.6%) | 0 (0.0%) | 5 (1.2%) | 4 (0.7%) |
| Native American/<br>American Indian/<br>Alaska Native | 0 (0.0%) | 0 (0.0%) | 1 (0.9%) | 4 (0.5%) | 0 (0.0%) | 0 (0.0%) | 1 (0.9%) | 0 (0.0%) | 1 (0.3%) | 0 (0.0%) | 3 (0.7%) | 2 (0.4%) |
| Native Hawaiian/<br>Pacific Islander | 0 (0.0%) | 0 (0.0%) | 0 (0.0%) | 1 (0.1%) | 0 (0.0%) | 0 (0.0%) | 0 (0.0%) | 0 (0.0%) | 0 (0.0%) | 0 (0.0%) | 0 (0.0%) | 0 (0.0%) |
| Multiracial | 1 (3.3%) | 0 (0.0%) | 3 (2.8%) | 22 (3.0%) | 0 (0.0%) | 0 (0.0%) | 5 (4.3%) | 9 (3.5%) | 8 (2.5%) | 0 (0.0%) | 22 (5.4%) | 15 (2.8%) |
| Other | 0 (0.0%) | 1 (14%) | 2 (1.9%) | 21 (2.8%) | 2 (6.5%) | 0 (0.0%) | 3 (2.6%) | 6 (2.3%) | 4 (1.2%) | 0 (0.0%) | 5 (1.2%) | 5 (0.9%) |
| **Region** |  |  |  |  |  |  |  |  |  |  |  |  |
| Midwest | 9 (30%) | 1 (13%) | 29 (26%) | 117 (16%) | 8 (26%) | 2 (29%) | 14 (12%) | 31 (12%) | 78 (24%) | 8 (17%) | 77 (19%) | 80 (15%) |
| Northeast | 7 (23%) | 3 (38%) | 22 (20%) | 176 (23%) | 5 (16%) | 1 (14%) | 22 (19%) | 32 (12%) | 63 (20%) | 9 (19%) | 72 (18%) | 70 (13%) |
| South | 7 (23%) | 3 (38%) | 35 (32%) | 277 (37%) | 15 (48%) | 3 (43%) | 67 (57%) | 156 (60%) | 131 (41%) | 22 (46%) | 190 (47%) | 283 (52%) |
| West | 7 (23%) | 1 (13%) | 25 (23%) | 277 (37%) | 3 (9.7%) | 1 (14%) | 14 (12%) | 40 (15%) | 49 (15%) | 9 (19%) | 68 (17%) | 110 (20%) |
| **Education** |  |  |  |  |  |  |  |  |  |  |  |  |
| High school or<br>less | 5 (17%) | 1 (14%) | 10 (9.0%) | 18 (2.4%) | 0 (0.0%) | 1 (14%) | 2 (1.7%) | 2 (0.8%) | 31 (9.7%) | 1 (2.1%) | 17 (4.2%) | 14 (2.6%) |
| Some college | 9 (30%) | 3 (43%) | 20 (18%) | 81 (11%) | 6 (19%) | 2 (29%) | 18 (15%) | 17 (6.6%) | 126 (39%) | 19 (40%) | 98 (24%) | 66 (12%) |
| Bachelor's degree | 8 (27%) | 1 (14%) | 37 (33%) | 222 (30%) | 10 (32%) | 3 (43%) | 35 (30%) | 68 (26%) | 77 (24%) | 10 (21%) | 119 (29%) | 144 (27%) |
| Any post-graduate<br>studies | 8 (27%) | 2 (29%) | 44 (40%) | 430 (57%) | 15 (48%) | 1 (14%) | 62 (53%) | 172 (66%) | 87 (27%) | 18 (38%) | 174 (43%) | 319 (59%) |

|  | Never-Infected<br>(n = 900) |  |  |  | Recovered-COVID<br>(n = 414) |  |  |  | Long-COVID<br>(n = 1,320) |  |  |  |
| --- | --- | --- | --- | --- | --- | --- | --- | --- | --- | --- | --- | --- |
|  | Unvaccinated<br>(n = 30) | Partially<br>Vaccinated<br>(n = 8) | Fully<br>Vaccinated<br>(n = 111) | Boosted<br>(n = 751) | Unvaccinated<br>(n = 31) | Partially<br>Vaccinated<br>(n = 7) | Fully<br>Vaccinated<br>(n = 117) | Boosted<br>(n = 259) | Unvaccinated<br>(n = 321) | Partially<br>Vaccinated<br>(n = 48) | Fully<br>Vaccinated<br>(n = 408) | Boosted<br>(n = 543) |
| **Pre-existing conditions** |  |  |  |  |  |  |  |  |  |  |  |  |
| Chronic lung disease | 0 (0.0%) | 0 (0.0%) | 3 (2.7%) | 8 (1.1%) | 1 (3.2%) | 1 (14%) | 0 (0.0%) | 1 (0.4%) | 7 (2.2%) | 1 (2.1%) | 4 (1.0%) | 8 (1.5%) |
| Asthma/reactive airway disease | 2 (6.7%) | 1 (13%) | 12 (11%) | 80 (11%) | 3 (9.7%) | 1 (14%) | 9 (7.7%) | 28 (11%) | 59 (18%) | 1 (2.1%) | 74 (18%) | 125 (23%) |
| Diabetes | 4 (13%) | 1 (13%) | 2 (1.8%) | 25 (3.3%) | 2 (6.5%) | 0 (0.0%) | 3 (2.6%) | 10 (3.9%) | 17 (5.3%) | 3 (6.2%) | 16 (3.9%) | 23 (4.2%) |
| History of stroke | 1 (3.3%) | 0 (0.0%) | 0 (0.0%) | 8 (1.1%) | 0 (0.0%) | 0 (0.0%) | 0 (0.0%) | 1 (0.4%) | 7 (2.2%) | 0 (0.0%) | 3 (0.7%) | 4 (0.7%) |
| Anxiety/other mental health condition | 6 (20%) | 4 (50%) | 32 (29%) | 237 (32%) | 7 (23%) | 3 (43%) | 22 (19%) | 64 (25%) | 93 (29%) | 16 (33%) | 124 (30%) | 201 (37%) |
| Depression | 3 (10%) | 3 (38%) | 24 (22%) | 159 (21%) | 5 (16%) | 0 (0.0%) | 17 (15%) | 40 (15%) | 80 (25%) | 15 (31%) | 99 (24%) | 153 (28%) |
| Cardiovascular disease<sup>a</sup> | 1 (3.3%) | 0 (0.0%) | 6 (5.4%) | 22 (2.9%) | 3 (9.7%) | 0 (0.0%) | 1 (0.9%) | 7 (2.7%) | 10 (3.1%) | 1 (2.1%) | 9 (2.2%) | 25 (4.6%) |
| Chronic kidney disease | 0 (0.0%) | 0 (0.0%) | 1 (0.9%) | 4 (0.5%) | 0 (0.0%) | 0 (0.0%) | 1 (0.9%) | 4 (1.5%) | 5 (1.6%) | 0 (0.0%) | 4 (1.0%) | 8 (1.5%) |
| Overweight/obesity | 17 (57%) | 6 (75%) | 64 (58%) | 406 (54%) | 19 (61%) | 6 (86%) | 63 (55%) | 127 (49%) | 219 (69%) | 32 (67%) | 258 (64%) | 324 (60%) |
| **SARS-CoV-2 variant<sup>b</sup>** |  |  |  |  |  |  |  |  |  |  |  |  |
| Alpha | - | - | - | - | 8 (26%) | 4 (57%) | 2 (1.7%) | 0 (0.0%) | 134 (42%) | 22 (45%) | 18 (4.4%) | 0 (0.0%) |
| Delta | - | - | - | - | 7 (23%) | 1 (14%) | 49 (42%) | 1 (0.4%) | 121 (38%) | 11 (23%) | 216 (53%) | 13 (2.4%) |
| Omicron | - | - | - | - | 16 (52%) | 2 (29%) | 66 (56%) | 258 (99%) | 66 (21%) | 15 (31%) | 174 (43%) | 530 (98%) |
| **SARS-CoV-2 vaccine type<sup>c</sup>** |  |  |  |  |  |  |  |  |  |  |  |  |
| mRNA | - | 8 (100%) | 109 (98%) | 704 (94%) | - | 7 (100%) | 108 (92%) | 245 (95%) | - | 48 (100%) | 366 (90%) | 513 (94%) |
| Adenovirus-vectored | - | 0 (0.0%) | 2 (1.8%) | 40 (5.3%) | - | 0 (0.0%) | 7 (6.0%) | 11 (4.3%) | - | 0 (0.0%) | 39 (9.6%) | 29 (5.3%) |
| Protein subunit | - | 0 (0.0%) | 0 (0.0%) | 2 (0.3%) | - | 0 (0.0%) | 1 (0.9%) | 2 (0.8%) | - | 0 (0.0%) | 1 (0.3%) | 0 (0.0%) |
| Other or unknown | - | 0 (0.0%) | 0 (0.0%) | 5 (0.6%) | - | 0 (0.0%) | 1 (0.9%) | 1 (0.4%) | - | 0 (0.0%) | 2 (0.5%) | 1 (0.2%) |

Abbreviations: SD: standard deviation; LTC: long-term care; K-12: kindergarten through grade 12; IT: information technology
<sup>a</sup>Includes cardiovascular/heart disease, history of congestive heart failure, and/or history of heart attack
<sup>b</sup>These dates were approximated: Alpha (between February 1, 2021, and June 30, 2021); Delta (between July 1, 2021, and November 30, 2021); Omicron (December 1, 2021, onward) [26].
<sup>c</sup>Pfizer and Moderna were classified as mRNA, Johnson & Johnson and AstraZeneca as adenovirus-vectored, and Novavax, Anhui Zhifei Longcom, Covaxin, CoronaVac, and Covilo as other.

Comorbidities were common across the cohort: 59% reported overweight/obesity, 22% depression, 31% anxiety, and 15% asthma/reactive airway disease. Among long-COVID participants, nearly all boosted participants were infected when the Omicron variant was the dominant circulating strain in the US (98%), whereas unvaccinated participants were more likely to have been infected during the Alpha (42%) or Delta (38%) periods.

### WMFI score distribution by infection status

We compared the unadjusted distributions WMFI scores across infection-status groups (Fig. 2). Participants with long-COVID exhibited higher mental fatigue scores (median = 11.5, IQR = 5.3-20) compared to recovered-COVID (median = 2.5, IQR = 0.5-5.7) and never-infected (median = 5.0, IQR = 2-11) participants. The spread in the long-COVID group was also broader, indicating greater variability. A previous analysis looking at the distribution of WMFI scores across the same three groups found that scores were stable across age strata [25], suggesting that the observed differences are unlikely to be driven by age.

### Association between vaccination status and mental fatigue in quantile regressions

Among participants with long-COVID, receiving a booster dose prior to SARS-CoV-2 infection was associated with lower WMFI scores across the outcome distribution, indicating less mental fatigue (Table 2 and Fig. 3). Compared with unvaccinated participants, boosted participants had WMFI scores that were 1.5 points lower at the 25^th^ percentile (*p* = 0.02), 2.6 points lower at the median (*p* = 0.01), and 3.8 points lower at the 75^th^ percentile (*p* = 0.002), reflecting a consistent downward shift in the score distribution (Fig. 3). A similar trend was observed in fully vaccinated participants with long-COVID. These differences were smaller and did not reach statistical significance (Table 2).

**Table 2.** Association between pre-SARS-CoV-2 infection COVID-19 vaccination and Wood Mental Fatigue Inventory score across infection and long-COVID status (*n* = 2,634).

| WMFI Score Difference<sup>a,b,c</sup> |  |  |  |
| --- | --- | --- | --- |
| | Never-Infected<br>($n = 900$) | Recovered-COVID<br>($n = 414$) | Long-COVID<br>($n = 1,320$) |
| **Percentile**<br>**(95% CI)** |  |  |  |
| **25<sup>th</sup>** |  |  |  |
| Unvaccinated | Ref | Ref | Ref |
| Full | 0.6 (-3.3, 4.4) | 0.0 (-0.5, 0.5) | -1.0 (-2.4, 0.4) |
| Boosted | 0.0 (-3.6, 3.6) | 0.0 (-0.5, 0.5) | -1.5 (-2.7, -0.2) |
| **50<sup>th</sup>** |  |  |  |
| Unvaccinated | Ref | Ref | Ref |
| Full | -0.1 (-5.9, 5.8) | 0.4 (-1.4, 2.2) | -1.1 (-3.3, 1.0) |
| Boosted | -1.1 (-6.8, 4.6) | 0.1 (-1.5, 1.7) | -2.6 (-4.7 -0.6) |
| **75<sup>th</sup>** |  |  |  |
| Unvaccinated | Ref | Ref | Ref |
| Full | -0.3 (-9.9, 9.4) | 0.0 (-2.8, 2.9) | -1.0 (-3.5, 1.4) |
| Boosted | -3.7 (-12.7, 5.3) | -0.3 (-3.0, 2.4) | -3.8 (-6.3, -1.4) |
| **Mixed Effect**<br>**(95% CI)** |  |  |  |
| Unvaccinated | Ref | Ref | Ref |
| Full | -0.1 (-3.3, 3.1) | 0.4 (-1.5, 2.2) | -1.9 (-3.3, -0.6) |
| Boosted | -2.2 (-5.2, 0.8) | -0.1 (-1.8, 1.7) | -4.0 (-5.3, -2.6) |

Abbreviations: CI: confidence interval, WMFI: Wood Mental Fatigue Inventory, Ref: Reference.
<sup>a</sup>Estimates represent the difference in WMFI score between vaccinated (full, boosted) (score differences shown in table) and unvaccinated participants. Negative values indicate lower WMFI scores (less mental fatigue).
<sup>b</sup>Models adjusted for gender, age, race, education, region, occupation, overweight/obesity, diabetes, history of stroke, chronic lung disease, asthma/airway reactive disease, cardiovascular disease, depression, anxiety/other mental health condition, and time since infection centered at 84 days. Time since infection was only included in the models for participants with a history of infection (recovered-COVID and long-COVID).
<sup>c</sup>Partial vaccination participants were omitted due to small sample sizes.

**Fig. 3.**
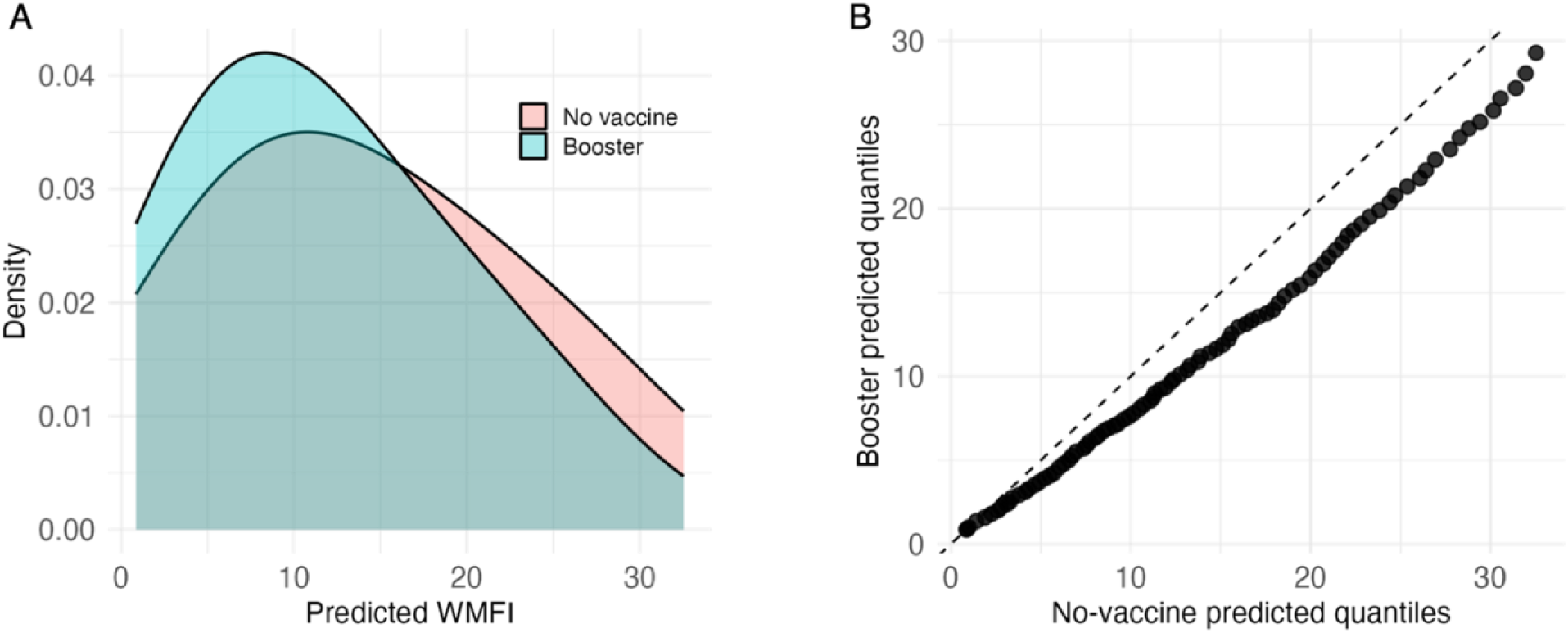
Estimated probability density function (A) and quantile-quantile plot (B) comparing vaccinated and unvaccinated participants with long-COVID at the time of infection.^a^. Abbreviations: WMFI: Wood Mental Fatigue Inventory ^a^Model adjusted for gender, age, race, education, region, occupation, overweight/obesity, diabetes, history of stroke, chronic lung disease, asthma/airway reactive disease, cardiovascular disease, depression, anxiety/other mental health condition, and time since infection centered at 84 days.

In contrast, vaccination status was not associated with WMFI scores at any percentile in the recovered-COVID or never-infected groups (negative controls). In both groups, point estimates were near 0, with wide confidence intervals observed in the never-infected group (Table 2). Due to the small number of partially vaccinated participants, we were underpowered to run quantile regression.

### Mixed-effects models

Using mixed-effects linear regression controlling for sociodemographic and clinical factors, boosted participants with long-COVID had WMFI scores that were, on average, 4 points lower than unvaccinated participants (*p* < 0.0001) (Table 2). Fully vaccinated participants with long-COVID also had lower mean WMFI scores compared to unvaccinated participants, consistent with a graded pattern in which booster vaccination is associated with the largest reduction. No significant difference was observed when evaluating the never-infected group (-2.2, *p* = 0.15) or the recovered-COVID group (-0.1, *p* = 0.92).

### Sensitivity Analyses

In a sensitivity analysis restricted to participants with long-COVID likely infected with the Omicron variant, the overall direction of association between booster vaccination and WMFI scores remained consistent with the main analysis, though effect sizes were no longer statistically significant, likely due to reduced sample size (Table S1). Across the outcome distribution, boosted participants had WMFI scores 1.1 points lower at the 25^th^ percentile (*p* = 0.5), 1.4 points lower at the 50^th^ percentile (*p* < 0.5), and 2.8 points lower at the 75^th^ percentile (*p* = 0.2) when compared to unvaccinated participants (*p* = 0.5). In the mixed-effects model, boosted participants had WMFI scores that were on average 2.4 points lower than their unvaccinated counterparts (*p* < 0.05).

## Discussion

In this analysis, we investigated the relationship between pre-infection COVID-19 vaccination and mental fatigue severity among participants with long-COVID in the JHCLS. The results demonstrated that participants with long-COVID who had been boosted prior to infection had lower mental fatigue scores compared to those with long-COVID who were unvaccinated, representing less mental fatigue. The reduction in mental fatigue appeared more pronounced at higher quantiles of the WMFI distribution, with a 1.5-point, 2.6-point, and 3.8-point reduction at the 25^th^, 50^th^, and 75^th^ percentiles, respectively.

To provide context to the magnitude of these associations, we compared unadjusted WMFI scores across groups. Participants with long-COVID displayed higher median WMFI scores than those with recovered-COVID (11 vs. 2), yielding a difference of 9 points. Thus, the 2.6-point reduction in median WMFI scores associated with being boosted pre-infection corresponds to approximately 29% of the difference between the long-COVID and recovered-COVID groups. Additionally, Bou-Holaigah et al. showed that WMFI score was strongly correlated with the 20-item Medical Outcomes Study (MOS-20) short form survey (*r* = -0.73; *p =* 0.01), which is a validated measure of general health [34]. Together, these observations suggest that the observed effect size is likely meaningful in terms of symptom burden. However, no established minimal clinically relevant difference exists for the WMFI in patients with long-COVID. Moreover, anchor-based calibrations linking WMFI scores to changes in functional outcomes are not yet available, so this comparison provides relative rather than absolute clinical context.

The overall pattern we observed suggests a dose-response relationship, with fully vaccinated participants showing lower WMFI scores than unvaccinated participants, although quantile estimates were smaller in magnitude and not statistically significant. Mixed effect models indicated lower mean WMFI in both fully vaccinated and boosted groups, with the largest difference among those who were boosted before infection. These associations may be confounded by the timing of infection relative to vaccine rollout and variant circulation, as fully vaccinated participants were more likely infected during earlier waves.

In sensitivity analyses restricted to participants infected during the Omicron period, quantile regression estimates remained directionally similar but were not statistically significant, likely reflecting reduced precision due to smaller sample size. However, the mixed-effects model estimate for boosted individuals remained statistically significant but attenuated. Other factors may also explain the attenuation of association. First, acute illness during Omicron waves appeared less severe than during Alpha or Delta waves, likely reflecting a combination of higher population immunity from vaccination and, to a lesser extent, possible differences in intrinsic virulence, though the relative contributions are difficult to disentangle [35–37]. Second, unrecognized prior SARS-CoV-2 infection prior to an Omicron-era infection may have generated partial immunity potentially diluting the marginal effect of a booster on post-acute symptom severity.

No association between vaccination status and WMFI scores was observed among recovered-COVID or never-infected participants. These groups were included as negative controls as our hypothesis was that long-COVID is a necessary component and any association among those that were never infected or recovered from their COVID infection would indicate bias (e.g., residual confounding, measurement error, or selection bias). In both negative control groups, vaccine was not associated with any change in mental fatigue. This pattern supports the specificity of the vaccination-WMFI association to individuals with long-COVID and argues against a spurious link driven by residual confounding.

Our findings align with prior reports showing that pre-infection vaccination is associated with reduced symptom severity among individuals with long-COVID [22,23]. Our findings differ from those of Mukherjee et al. [38] who found no association between pre-infection vaccination and the severity of neurological symptoms in long-COVID. Since mental fatigue and brain fog, which is considered a neurological symptom, are highly correlated [39], this discrepancy warrants further investigation. Mukherjee et al.’s study population consisted exclusively of patients referred to a tertiary neuro-COVID clinic, including both post-hospitalization and non-hospitalized individuals, most of whom reported persistent neurological symptoms such as brain fog, headache, and dizziness. In contrast, our study was conducted in a community-based, nationwide virtual cohort that included both participants with and without prior SARS-CoV-2 infection, representing a broader spectrum of long-COVID severity. Clinic-based samples often capture individuals with more severe or refractory symptoms and may differ in demographic and clinical composition from community cohorts, which could partly explain these divergent findings. Additionally, Mukherjee et al. relied on data collected from a single time point or within a narrow period of time, and did not adjust for potential confounders, further limiting comparability.

Our study has several notable strengths. First, the JHCLS is well-characterized, with detailed demographic, socioeconomic, and clinical data, including regional and comorbidity profiles, allowing for more comprehensive adjustments in the analysis. Second, the study employed a validated measure of mental fatigue, ensuring a standardized and reliable assessment of the outcome. Additionally, because data were collected remotely through a nationwide virtual cohort, participants who were too sick to travel to a clinic were still able to participate. This design improves the representativeness of the sample by including those who might otherwise be excluded from traditional in-person studies. Fourth, we included negative controls. The lack of an association between vaccination and WMFI among those who were never infected or recovered from their COVID infection supports that the relationship between vaccination and WMFI among those with long-COVID is not a spurious association.

This study also has limitations. Our sample over-represents participants who identify as women, white, and those from higher educational backgrounds, which limits generalizability if these factors are effect modifiers of the relationship between vaccination and WMFI among those with long-COVID [40].

## Conclusion

In this nationwide virtual cohort, receiving a COVID-19 booster dose prior to SARS-CoV-2 infection was associated with lower long-COVID–associated mental fatigue. Fully vaccinated participants also had lower WMFI scores compared to unvaccinated individuals, with smaller effects than those observed in boosted participants, consistent with a dose-response relationship. Associations were consistent across the distribution of scores and in mixed-effects models and were absent among recovered-COVID and never-infected controls, suggesting specificity to long-COVID.

As mental fatigue remains a common and disabling symptom of long-COVID, our results underscore the potential added value of vaccination in reducing long-term functional impairment. Future work should evaluate the biological and immunological mechanisms underlying this association, assess its durability over time, and determine whether additional booster doses further modify mental fatigue severity. Long-COVID remains a serious public health concern. Understanding how vaccination affects neurological symptoms like mental fatigue can inform future prevention strategies.

## Author contributions

Priya Duggal, Bryan Lau, Shruti H. Mehta, and Madeline Sagona conceptualized and designed the study. Priya Duggal, Bryan Lau, Shruti H. Mehta, Zhanmo Ni, and Eryka Wentz contributed to data curation. Madeline Sagona formulated the research question, performed the statistical analyses, and produced the tables and figures. Madeline Sagona conducted the literature review and wrote the original draft of the manuscript, and Bryan Lau and Eryka Wentz provided supervision. All authors provided critical review and revision of the manuscript.

## Acknowledgements

We would like to thank our participants for their time, energy, and support. Their input has been invaluable to our understanding of long-COVID. We would also like to thank our student researchers, Riya Karnik, Kristen Chen, and Yuzheng Xing.

## Conflict of interest statement

Shruti H. Mehta reports material support from Abbott Diagnostics unrelated to this study.

## Funding information

None

## Data availability statement

The data underlying this article will be shared upon reasonable request to the corresponding author.

## Notes

### Funding Statement

This study did not receive any funding

### Author Declarations

The Institutional Review Board of Johns Hopkins Bloomberg School of Public Health gave ethical approval for this work on December 15, 2020.

