## Supplementary Material for "Pre-infection COVID-19 vaccination and long-COVID mental fatigue severity: Findings from the Johns Hopkins COVID Long Study"

**Supporting Information**

**S Table 1**. Association between pre-SARS-CoV-2 infection COVID-19 vaccination and Wood Mental Fatigue Inventory score among participants with long-COVID who were likely infected with the Omicron variant (*n* = 785).

| | **Omicron only WMFI Score Difference**[a,b] |
|---|---|
| **Percentile (95% CI)** | |
| **25th** | |
| Unvaccinated | Ref |
| Full[c] | -0.5 (-3.2, 2.2) |
| Boosted | -1.1 (-3.4, 1.3) |
| **50th** | |
| Unvaccinated | Ref |
| Full | -0.2 (-5.3, 4.8) |
| Boosted | -1.4 (-6.0, 3.2) |
| **75th** | |
| Unvaccinated | Ref |
| Full | 0.7 (-4.5, 5.9) |
| Boosted | -2.8 (-7.4, 1.7) |
| **Mixed Effect (95% CI)** | |
| Unvaccinated | |
| Full | -0.4 (-2.9, 2.2) |
| Boosted | -2.4 (-4.7, -0.0) |

Abbreviations: WMFI: Wood Mental Fatigue Inventory Score, CI: confidence interval, Ref: Reference.

[a]Estimates represent the difference in WMFI score between vaccinated (partial, full, boosted) (score differences shown in table) and unvaccinated participants. Negative values indicate lower scores (less mental fatigue).

[b]Model adjusted for gender, age, race, education, region, occupation, overweight/obesity, diabetes, history of stroke, chronic lung disease, asthma/airway reactive disease, cardiovascular disease, depression, anxiety/other mental health condition, and time since infection centered at 84 days.

**S Table 2.** Occupational characteristics in the Johns Hopkins Covid Long Study (*n* = 2,634).

| | **Never-Infected** (*n* = 900) | | | | **Recovered-COVID** (*n* = 414) | | | | **Long-COVID** (*n* = 1,320) | | | |
|---|---|---|---|---|---|---|---|---|---|---|---|---|
| | Unvaccinated (*n* = 30) | Partially Vaccinated (*n* = 8) | Fully Vaccinated (*n* = 111) | Boosted (*n* = 751) | Unvaccinated (*n* = 31) | Partially Vaccinated (*n* = 7) | Fully Vaccinated (*n* = 117) | Boosted (*n* = 259) | Unvaccinated (*n* = 321) | Partially Vaccinated (*n* = 48) | Fully Vaccinated (*n* = 408) | Boosted (*n* = 543) |
| **Occupation** | | | | | | | | | | | | |
| Healthcare worker (hospital) | 0 (0.0%) | 0 (0.0%) | 5 (4.5%) | 47 (6.3%) | 1 (3.2%) | 0 (0.0%) | 28 (24%) | 32 (12%) | 15 (4.7%) | 2 (4.2%) | 43 (11%) | 42 (7.7%) |
| Healthcare worker (community) | 3 (10%) | 0 (0.0%) | 8 (7.2%) | 45 (6.0%) | 1 (3.2%) | 0 (0.0%) | 9 (7.7%) | 23 (8.9%) | 26 (8.1%) | 4 (8.3%) | 34 (8.3%) | 45 (8.3%) |
| Nursing home/ LTC or social service worker | 0 (0.0%) | 0 (0.0%) | 2 (1.8%) | 7 (0.9%) | 0 (0.0%) | 0 (0.0%) | 1 (0.9%) | 3 (1.2%) | 2 (0.6%) | 2 (4.2%) | 3 (0.7%) | 3 (0.6%) |
| Teacher (K-12)/ educator | 2 (6.7%) | 0 (0.0%) | 9 (8.1%) | 34 (4.5%) | 0 (0.0%) | 0 (0.0%) | 9 (7.7%) | 11 (4.2%) | 18 (5.6%) | 4 (8.3%) | 23 (5.6%) | 43 (7.9%) |
| Police/guard/ emergency services or military/national service | 1 (3.3%) | 0 (0.0%) | 0 (0.0%) | 3 (0.4%) | 0 (0.0%) | 0 (0.0%) | 0 (0.0%) | 2 (0.8%) | 0 (0.0%) | 1 (2.1%) | 2 (0.5%) | 8 (1.5%) |
| Transportation or laborer | 1 (3.3%) | 0 (0.0%) | 1 (0.9%) | 2 (0.3%) | 0 (0.0%) | 0 (0.0%) | 0 (0.0%) | 0 (0.0%) | 3 (0.9%) | 0 (0.0%) | 2 (0.5%) | 2 (0.4%) |
| Hotel/hospitality staff or cleaning | 1 (3.3%) | 0 (0.0%) | 4 (3.6%) | 3 (0.4%) | 0 (0.0%) | 0 (0.0%) | 1 (0.9%) | 0 (0.0%) | 3 (0.9%) | 0 (0.0%) | 0 (0.0%) | 1 (0.2%) |
| Bartender/ restaurant worker or food prep | 3 (10%) | 0 (0.0%) | 0 (0.0%) | 8 (1.1%) | 0 (0.0%) | 0 (0.0%) | 1 (0.9%) | 2 (0.8%) | 4 (1.2%) | 0 (0.0%) | 2 (0.5%) | 6 (1.1%) |
| Sales, engineer, researcher/ scientist, legal/ government worker, or computer/IT | 1 (3.3%) | 0 (0.0%) | 1 (0.9%) | 16 (2.1%) | 0 (0.0%) | 0 (0.0%) | 2 (1.7%) | 9 (3.5%) | 3 (0.9%) | 1 (2.1%) | 7 (1.7%) | 19 (3.5%) |
| Other | 18 (60%) | 8 (100%) | 81 (73%) | 586 (78%) | 29 (94%) | 7 (100%) | 66 (56%) | 177 (68%) | 247 (77%) | 34 (71%) | 292 (72%) | 374 (69%) |

**Figure S1.** Directed acyclic graph of the relationships between vaccination, potential confounders, COVID-19, long-COVID, and mental fatigue.

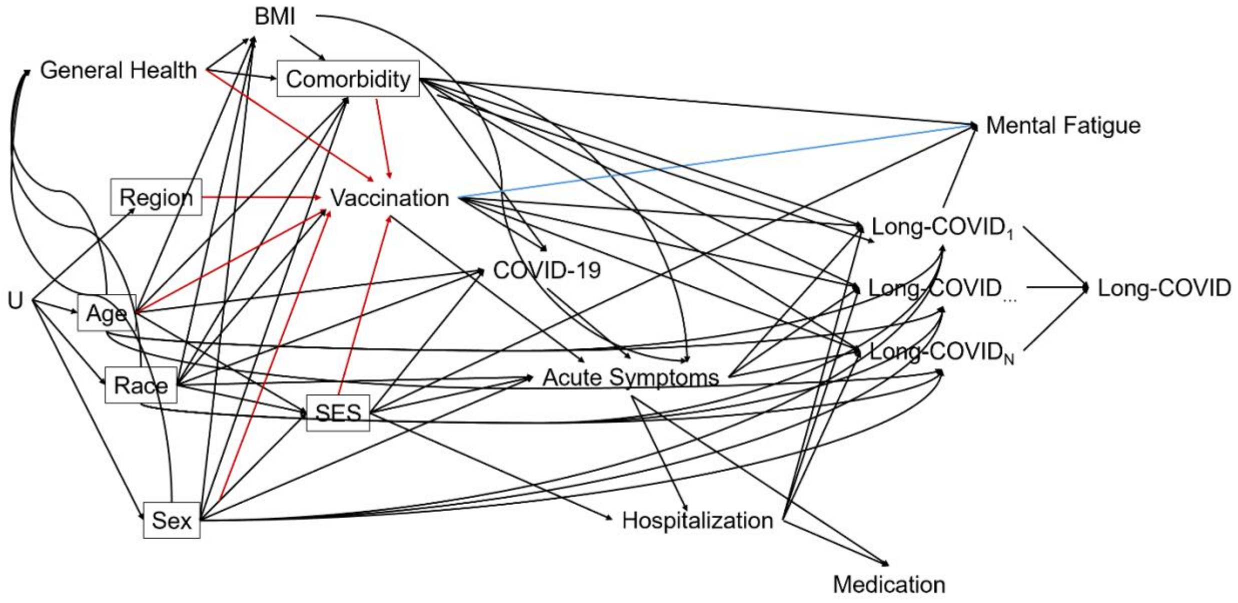


Abbreviations: BMI: body mass index, SES: socioeconomic status

Figure S1 shows the directed acyclic graph (DAG) of the model that guided these analyses. We assumed that the covariates of age, race, gender, region, socioeconomic status (SES) as measured by occupation, and the presence of comorbid conditions were the pertinent confounders that needed to be accounted for in order to block the non-causal pathways between vaccination and mental fatigue. We further assumed that the relationship between vaccination and mental fatigue only existed in the presence of long-COVID. That is, we assumed that vaccination's effect on mental fatigue was only biologically plausible when participants had long-COVID and that vaccination has no effect among those who were never infected and among those who recovered from their SARS-CoV-2 infection without developing long-COVID. Factors that do impact mental fatigue included other comorbidities and occupation which was measured in part for SES. Note, we split long-COVID into long-COVID$_1$, ..., long-COVID$_N$ in order to show different potential mechanisms that have been posited (e.g., viral remnants, immune dysfunction, etc.). However, since we do not know how to identify the mechanism, we create an observable general long-COVID that can be identified to restrict the sample for the nested samples (Figure S2
